# The Clinical Utility of Serial Procalcitonin and Procalcitonin Clearance in Predicting the Outcome of COVID-19 Patients

**DOI:** 10.1101/2021.06.14.21258855

**Authors:** Sara I. Taha, Aalaa K. Shata, Shereen A. Baioumy, Shaimaa H. Fouad, Aya H. Moussa, Mariam K. Youssef

**Author notes:** **Corresponding Author:** Sara Ibrahim Taha, MD, PhD, Lecturer of Clinical Pathology/ Immunology, Faculty of Medicine, Ain Shams University., Address: Ain Shams University, Abassia, Cairo, Egypt.

## Abstract

**Background:** The pandemic of coronavirus disease 2019 (COVID-19) represents a significant threat to global health. Sensitive tests that effectively predict the disease outcome are essentially required to guide proper intervention.

**Objectives:** To evaluate the predictive ability of serial procalcitonin (PCT) measurement to predict the outcome of COVID-19 patients, using PCT clearance (PCT-c) as a tool to reflect its dynamic changes.

**Methods:** A prospective observational study of inpatients diagnosed with COVID-19 at the Quarantine Hospitals of Ain-Shams University, Cairo, Egypt. During the first five days of hospitalization, serial PCT and PCT-c values were obtained and compared between survivors and non-survivors. Patients were followed up to hospital discharge or in-hospital mortality.

**Results:** Compared to survivors, serial PCT levels of non-survivors were significantly higher (p ≤ 0.001) and progressively increased during follow-up. In contrast, PCT-c values were significantly lower (p < 0.01) and progressively decreased. Receiver operating characteristic (ROC) curve analysis showed that using the initial PCT value alone, at a cut-off value of 0.80 ng/ml, the area under the curve for predicting in-hospital mortality was 0.81 with 61.1% sensitivity and 87.3% accuracy. Serial measurements showed better predictive performance, and the combined prediction value was better than the single prediction by the initial PCT alone.

**Conclusions:** Serial PCT measurement could be a helpful laboratory tool to predict the prognosis and outcome of COVID-19 patients. Moreover, PCT-c could be a reliable tool to assess PCT progressive kinetics.

## INTRODUCTION

Coronavirus disease 2019 (COVID-19) is highly infectious pneumonia caused by severe acute respiratory syndrome coronavirus 2 (SARS-CoV-2). It first emerged in December 2019 in Wuhan, China, then rapidly spread worldwide, and now it is considered a pandemic that represents an urgent threat to global health.^**1**^ Fever, dry cough, and fatigue are the main manifestations of COVID-19. In the more severe cases, patients may have dyspnea and hypoxia five to seven days after the onset of symptoms that can rapidly progress to respiratory distress, coagulopathy, multi-organ failure, and even death.^**2**^ The mortality of COVID-19 pneumonia is higher than other viral pneumonia.^**3**^ Since inflammation is an important factor in COVID-19 mortality, sensitive inflammatory biomarkers that reflect lung lesion changes should be continuously explored.^**1**^

Procalcitonin (PCT) is a calcitonin-related pro-hormone released by the thyroid parafollicular cells. Under physiological conditions, serum PCT is usually below 0.05 ng/ml. However, its levels increase significantly 2–6 hours after stimulation by microbial infection, being released by all parenchymal tissues under the effect of endotoxins and pro-inflammatory cytokines.^**4**^ Its role in inflammatory response includes chemotactic function, modulation of inducible nitric oxide synthase, and induction of cytokines. So, it has been used as a marker of prognosis in sepsis.^**5**^

Our study aimed to explore the ability of serial serum concentrations and clearance of PCT to predict the prognosis and mortality of COVID-19 patients.

## Methodology

### Study design and patient selection

This prospective observational study included 63 adults (age ≥ 18 years) of COVID-19 patients admitted to the Quarantine Hospitals of Ain-Shams University (El-demerdash Hospital and Field Hospital) in Cairo, Egypt, from April 10 to May 15, 2021. All included patients needed to have 1) laboratory-confirmed SARS-CoV-2 infection by a reverse transcription-PCR test from a nasopharyngeal swab prior to hospitalization; and 2) at least one PCT measurement upon hospital admission (or within 24 hours) in their medical records. Pregnant women and patients with non-infectious causes of systemic inflammation that can induce PCT production, such as trauma, surgery, burn injury, and chronic kidney disease, were excluded. In this study, patients were followed up to hospital discharge or in-hospital mortality. Patients’ severity categorization, treatment, and discharge criteria followed the management protocol of COVID-19 patients released by Ain Shams University Hospitals.^**6**^

### Data collection

For all included patients, baseline and follow-up data were collected from medical records and encompassed demographics, clinical history, presence of comorbidities, routine laboratory test findings, length of hospital stay, ICU admission, and clinical outcome.

### Procalcitonin (PCT) measurement

Because of the difficult access to some patients with critical conditions, in addition to the death or hospital discharge of others, not all initially included patients were subjected to serial PCT evaluation. Initial PCT values were obtained from the medical records of all included patients (n=63). Then, further assessment of PCT levels was done using serum samples that were collected on hospital days 3 (n = 47) and 5 (n = 49), and were stored at −80°C until analysis, using the Elecsys® BRAHMS PCT sandwich immunoassay principle of the electrochemiluminescence autoanalyzer (COBAS e411; Roche Diagnostics GmbH, Mannheim, Germany) according to the manufacturer’s instructions, with the analytical measuring range of 0.02-100 ng/ml and the detection limit of < 0.02 ng/ml. Initial PCT that was done upon hospital admission or within 24 hours and obtained from patients’ medical records was defined as PCTD1, while PCT values that were measured on day 3 (48-72 hours) and day 5 (96 – 120 hours) were defined as PCTD3 and PCTD5, respectively.

### Procalcitonin clearance (PCT-c) calculation

The clearance of procalcitonin (PCT-c) on days 3 (PCT-cD3) and 5 (PCTc-D5) was calculated as follows: PCT-c D3or5 (%) = [(PCTD1 – PCTD3or5)/PCTD1] × 100. Clearance values are positive with decreasing concentrations and negative with increasing concentrations.^**7**^

### Statistical methods

The IBM SPSS statistics program (V. 26.0, IBM Corp., USA, 2019) was used for data analysis. Quantitative non-parametric measures were expressed as medians and percentiles. In addition, categorized data were expressed in the form of numbers and percentages. Comparison between two independent groups for non-parametric data was done using the Wilcoxon Rank Sum test. Meanwhile, the Chi-square test was used to compare the categorized data. A diagnostic validity test was and the area under the curve (AUC) was calculated after the receiver operating characteristic (ROC) curve was constructed to determine each variable’s most sensitive and specific predictive cutoff point. All statistical tests were two-tailed, and the probability of error at 0.05 was considered significant. Data points that were missing were not extrapolated.

## Results

### Demographic and clinical characteristics

During the enrollment period of this study, a final sample of 63 patients, who met the inclusion criteria, was initially obtained. The median age was 55 years (IQR:43– 70), and 54.0% of patients were men. Hypertension (38.1%) and diabetes (30.2%) were the most common comorbidities. Regarding admission disease severity, 50.8% (n=32) of the included patients had a non-severe disease, while 49.2% (n=31) were severe. The median length of stay in the hospital was nine days (IQR: 7 -12) and ranged from 4 to 30 days. In total, 29 patients (46.0%) were admitted to the ICU, and 18 (28.6%) died.

The characteristics of non-survivors were similar to survivors regarding age, gender, length of hospital stay, and underlying comorbidities, except for diabetes mellitus, which was significantly associated with in-hospital mortality (P = 0.001). Similarly, all non-survivors (n = 18) had severe disease, and 94.4% (n = 17) were admitted to the ICU. In comparison, only 28.9% (n = 13) of survivors were considered severe, and 26.7% (n = 12) required ICU admission. The demographic and clinical characteristics of the included patients are presented in **table 1**.

**Table 1:** Demographic and clinical characteristics of the studied population

| Variable |  | All Cases<br>(n=63) | Survivors<br>(n=45) | Non– survivors<br>(n=18) | p–<br>value |
| --- | --- | --- | --- | --- | --- |
| Age (years) | Median (IQR) | 55 (43– 70) | 54 (37.5– 66) | 63.5 (52.25– 73) | 0.052 |
|  | Range | (21– 94) | (21– 94) | (42– 85) |  |
| Sex n, (%) | Male | 34 (54.0%) | 24 (53.3%) | 10 (55.6%) | 0.873 |
|  | Female | 29 (46.0%) | 21 (46.7%) | 8 (44.4%) |  |
| Comorbidities<br>n, (%) | DM | 19 (30.2%) | 8 (17.8%) | 11 (61.1%) | **0.001** |
|  | HTN | 24 (38.1%) | 14 (31.1%) | 10 (55.6%) | 0.071 |
|  | COPD | 9 (14.3%) | 5 (11.1%) | 4 (22.2%) | 0.255 |
|  | IHD | 10 (15.9%) | 6 (13.3%) | 4 (22.2%) | 0.383 |
| Severity n, (%) | Non– severe | 32 (50.8%) | 32 (71.1%) | 0 (0.0%) | **≤ 0.001** |
|  | Severe | 31 (49.2%) | 13 (28.9%) | 18 (100.0%) |  |
| Hospital stay<br>(days) | Median (IQR) | 9 (7– 12) | 9 (6.5– 12) | 11(8– 19.25) | 0.051 |
|  | Range | (4– 30) | (4– 23) | (4– 30) |  |
| ICU admission<br>n, (%) | Negative | 34 (54.0%) | 33 (73.3%) | 1 (5.6%) | **≤ 0.001** |
|  | Positive | 29 (46.0%) | 12 (26.7%) | 17 (94.4%) |  |

DM: diabetes mellitus; HTN: hypertension; COPD: chronic obstructive pulmonary disease; IHD: ischemic heart disease; ICU: intensive care unit; IQR: interquartile range. Statistical significance set at 0.05.

### Prognostic value of serial PCT and PCT-c

**Table 2** presents the values of PCT and PCT-c with their serial changes in survivors and non-survivors. Serum levels of PCT increased significantly with in-hospital mortality. Compared with survivors, non-survivors showed significantly higher values of PCTD1 (median: 1.20 ng/ml (IQR:0.23 – 1.96) vs. 0.12 ng/ml (0.06 – 0.34); P ≤ 0.001), PCTD3 (median: 1.6 ng/ml (0.99 – 2.2) vs. 0.08 ng/ml (0.06 – 0.14); P ≤ 0.001) and PCTD5 (median: 3.2 ng/ml (IQR: 2.05 – 7.1) vs. 0.05 ng/ml (0.04 – 0.08); P ≤ 0.001).

**Table 2:**
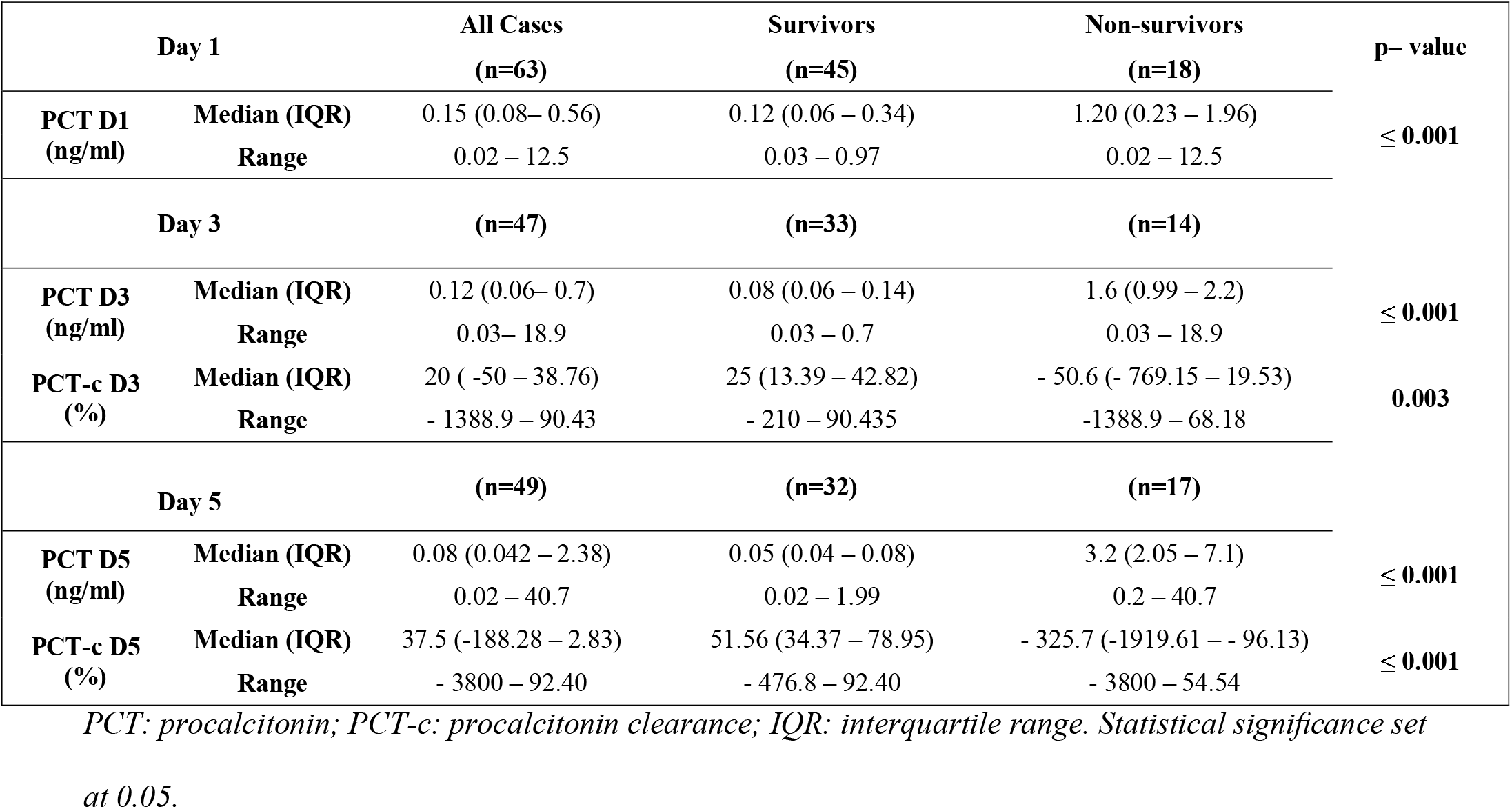
Comparisons of values of PCT and PCT-c, and their serial changes between survivors and non-survivors

| Day 1 |  | All Cases<br>(n=63) | Survivors<br>(n=45) | Non-survivors<br>(n=18) | p- value |
| --- | --- | --- | --- | --- | --- |
| **PCT D1**<br>(ng/ml) | **Median (IQR)** | 0.15 (0.08– 0.56) | 0.12 (0.06 – 0.34) | 1.20 (0.23 – 1.96) | **≤ 0.001** |
|  | **Range** | 0.02 – 12.5 | 0.03 – 0.97 | 0.02 – 12.5 |  |
| Day 3 |  | (n=47) | (n=33) | (n=14) |  |
| **PCT D3**<br>(ng/ml) | **Median (IQR)** | 0.12 (0.06– 0.7) | 0.08 (0.06 – 0.14) | 1.6 (0.99 – 2.2) | **≤ 0.001** |
|  | **Range** | 0.03– 18.9 | 0.03 – 0.7 | 0.03 – 18.9 |  |
| **PCT-c D3**<br>(%) | **Median (IQR)** | 20 ( -50 – 38.76) | 25 (13.39 – 42.82) | - 50.6 ( - 769.15 – 19.53) | **0.003** |
|  | **Range** | - 1388.9 – 90.43 | - 210 – 90.435 | -1388.9 – 68.18 |  |
| Day 5 |  | (n=49) | (n=32) | (n=17) |  |
| **PCT D5**<br>(ng/ml) | **Median (IQR)** | 0.08 (0.042 – 2.38) | 0.05 (0.04 – 0.08) | 3.2 (2.05 – 7.1) | **≤ 0.001** |
|  | **Range** | 0.02 – 40.7 | 0.02 – 1.99 | 0.2 – 40.7 |  |
| **PCT-c D5**<br>(%) | **Median (IQR)** | 37.5 (-188.28 – 2.83) | 51.56 (34.37 – 78.95) | - 325.7 (-1919.61 – - 96.13) | **≤ 0.001** |
|  | **Range** | - 3800 – 92.40 | - 476.8 – 92.40 | - 3800 – 54.54 |  |

*PCT: procalcitonin; PCT-c: procalcitonin clearance; IQR: interquartile range. Statistical significance set at 0.05.*

As regards PCT-c, it decreased significantly with in-hospital mortality. Values of PCT-cD3 and PCT-cD5 were significantly lower in non-survivors compared to survivors, (median: □50.6% (IQR: □ 769.15 – 19.53) vs. 25% (13.39 – 42.82); P = 0.003 and median: □ 325.7% (IQR □ 1919.61 – □ 96.13) vs. 51.56 % (34.37 – 78.95); P ≤ 0.001, respectively).

In non-survivors and during the follow-up, serial PCT levels showed an overall progressive increase. In contrast, PCT-c values progressively decreased. **Figures 1 and 2**

**Figure 1:**
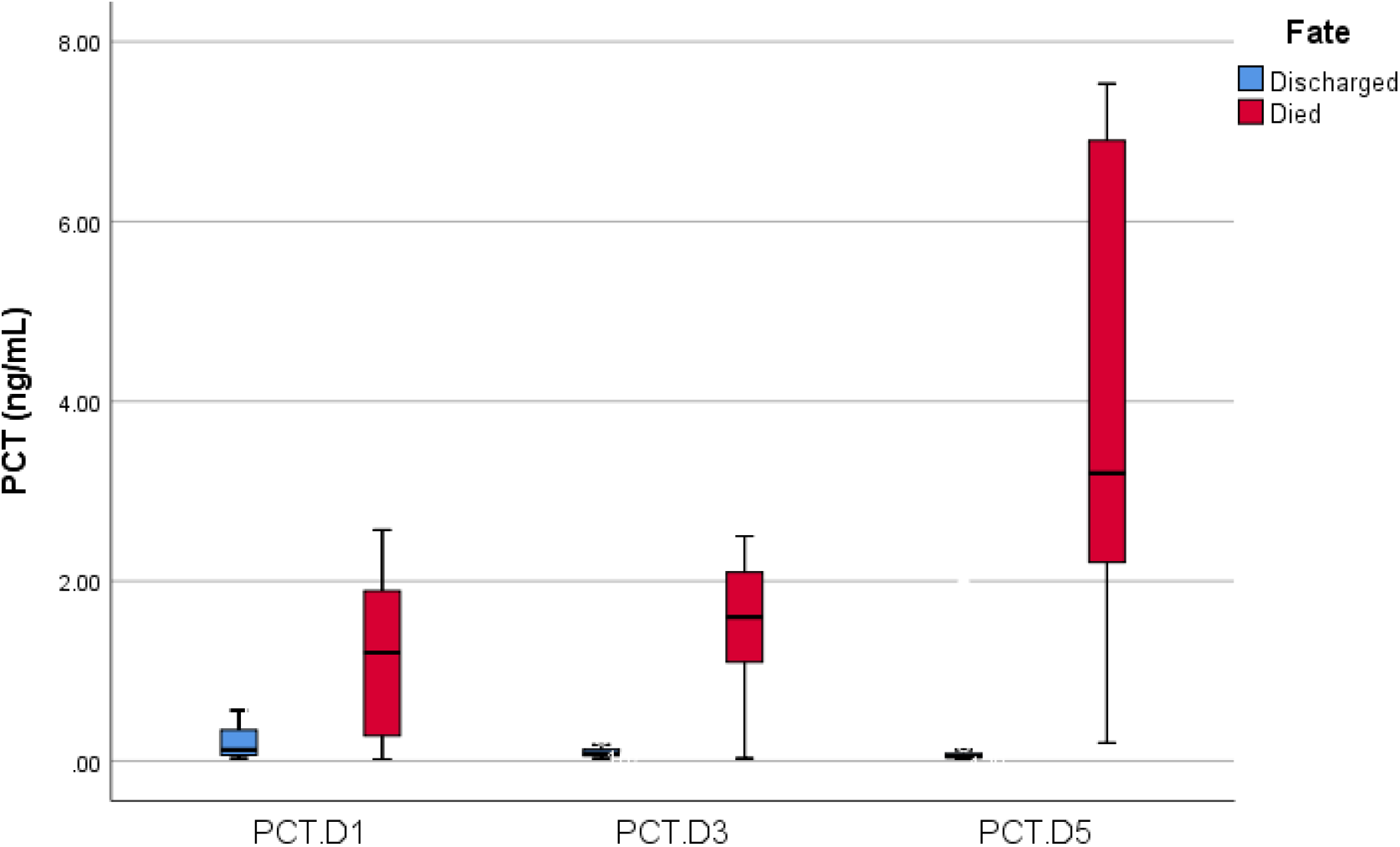
Box plot representing the range, median, and quartiles of serial PCT values on hospital days 1, 3 and 5 in survivors and non-survivors (tested by Wilcoxon rank-sum test)

**Figure 2:**
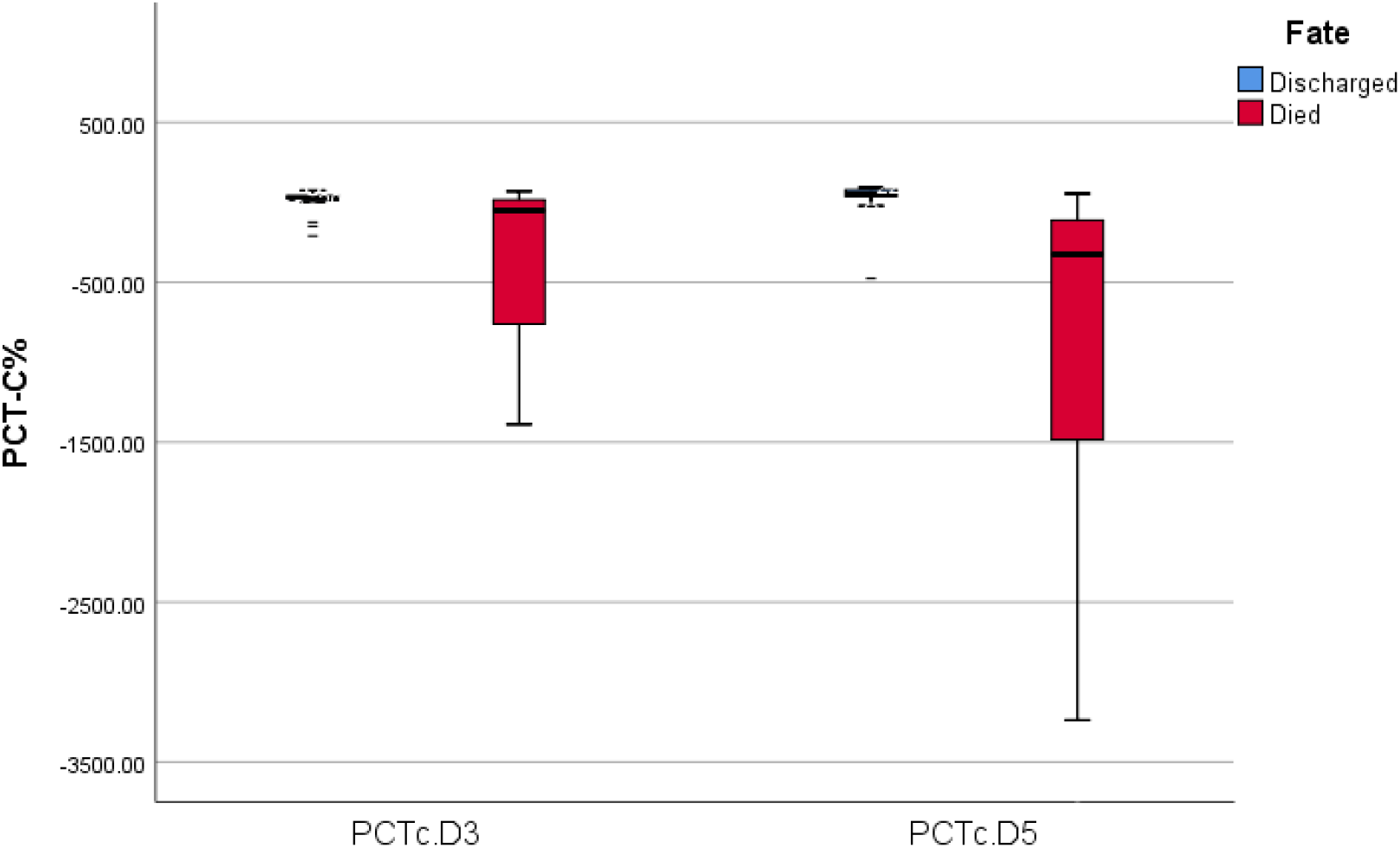
Box plot representing the range, median, and quartiles of PCT-c values on hospital days 3 and 5 in survivors and non-survivors. (tested by Wilcoxon rank-sum test)

**Table 3** demonstrates the diagnostic validity and ROC curve analysis of the ability of serial PCT and PCT-c to predict in-hospital mortality of COVID-19 patients. Regarding the single indicators, serial PCT and PCT-c showed an overall better predictive performance for in-hospital mortality than the initial PCT measurement. At the optimum cutoff values of 0.80 ng/ml, 0.54 ng/ml, 0.18 ng/ml, - 11.1 % and -79.77% for PCTD1, PCTD3, PCTD5, PCT-cD3 and PCT-cD5, respectively, the AUC followed the order of PCTD5> PCT-c D5>PCTD3>PCTD1> PCT-c D3 being 0.96, 0.89, 0.82, 0.81 and 0.74, respectively. Despite PCTD1 had the highest diagnostic specificity (97.8%), it showed the lowest diagnostic sensitivity (61.1%). PCTD5 then PCTD3 had the highest diagnostic sensitivity (100% and 85.7%, respectively), NPV (100% and 94.1%, respectively) and diagnostic accuracy (95.9% and 93.6%, respectively). The best PPV was of PCT-c D5 then PCTD3 (93.3% and 92.3%, respectively). PCT-c D3 showed the lowest specificity (87.9%), PPV (71.4%) and accuracy (83.0%).

**Table 3:** Diagnostic validity test and receiver operating characteristic (ROC) analysis of the ability of serial PCT measurements and PCT-c to predict in-hospital mortality of COVID-19 patients

| Variable | Cut off | AUC | Specificity (%) | Sensitivity (%) | NPV (%) | PPV (%) | Accuracy (%) |
| --- | --- | --- | --- | --- | --- | --- | --- |
| **PCTD1 (ng/ml)** | 0.80 | 0.81 | 97.8 | 61.1 | 86.3 | 91.7 | 87.3 |
| **PCTD3 (ng/ml)** | 0.54 | 0.82 | 97.0 | 85.7 | 94.1 | 92.3 | 93.6 |
| **PCTD5 (ng/ml)** | 0.18 | 0.96 | 93.8 | 100.0 | 100.0 | 89.5 | 95.9 |
| **PCT-c D3 (%)** | - 11.1 | 0.74 | 87.9 | 71.4 | 87.9 | 71.4 | 83.0 |
| **PCT-c D5 (%)** | -79.77 | 0.89 | 96.9 | 82.4 | 91.2 | 93.3 | 91.8 |
| **PCTD1 at 0.80 + PCTD3 at 0.03** | -- | 0.93 | 97.0 | 92.9 | 97.0 | 92.9 | 95.7 |
| **PCTD1 at 0.80 + PCTD5 at 0.398** | -- | 0.98 | 100.0 | 94.1 | 97.0 | 100.0 | 98.0 |
| **PCTD1 at 0.80 + PCT-cD3 at-50** | -- | 0.96 | 100.0 | 92.9 | 97.1 | 100.0 | 97.9 |
| **PCTD1 at 0.80 + PCT-cD5 at 54.54** | -- | 1.00 | 100.0 | 100.0 | 100.0 | 100.0 | 100.0 |

*PPV: positive predictive value; NPV: negative predictive value; AUC: area under curve; PCT: procalcitonin; PCT-c: procalcitonin clearance.*

The AUC increased when PCTD1 at a cut off of 0.8 ng/ml was added to either PCTD3 at 0.03 ng/ml, PCTD5 at 0.398 ng/ml, PCT-cD3 at-50%, or PCT-cD5 at 54.54% to be 0.93, 0.98, 0.96, and 1.00, respectively, showing that the combined prediction value was better than the single prediction by initial PCT value, whereas, the best predictive performance was seen in the PCTD1+PCTD-c5 combination, having 100% specificity, sensitivity, PPV, NPV, and accuracy in predicting COVID-19 patients’ in-hospital mortality.

## Discussion

The COVID19 pandemic has had a significant influence on global health. In many nations, the fast spread of the virus, high caseloads, and high proportions of patients requiring hospitalization, ICU admission, and respiratory support have put unparalleled demand on healthcare facilities and manpower.^**3**^ It is vital to implement sensitive testing that effectively predicts prognosis to guide optimal intervention to reduce hospital stay and in-hospital mortality. ^**5**^ Since 1993, procalcitonin has been a prognostic inflammatory biomarker for determining the severity of infectious etiologies.^**8**^ As a result, several studies have found a significant increase in serum PCT levels after severe lung bacterial infections, since cytokines directly trigger its release. PCT levels were normal or slightly raised during viral infections and non-specific inflammatory processes, owing to TNF**-**α suppression by virus-stimulated interferon**-**γ synthesis (INF**-**γ). ^**9**,**10**^ Despite being a highly utilized biomarker of bacterial infection, contrasting opinions exist regarding its efficacy in predicting the prognosis of COVID-19 patients.^**11**,**12**^ However, in severe COVID-19 infections, some studies found elevated levels of PCT^**13**,**14**^, while others found normal levels.^**15**,**16**^

To further understand this association, we conducted this study, in which the PCT level was determined sequentially in adult Egyptian COVID-19 patients during the first five days of hospital admission to study PCT kinetics and prognostic performance through the course of the disease. By following up on the patients, we found that the PCT level was significantly higher (p ≤ 0.001) in non-survivors than survivors with an overall progressive increasing tendency, proposing it a reliable prognostic biomarker of in-hospital mortality of COVID-19 patients.

Consistent with our findings, in a study by Xu et al., increased serum PCT was an independent risk factor for mortality in hospitalized COVID-19 patients.^**1**^ Similarly, a four-study pooled analysis found that greater PCT was linked to five-fold increased risk of severe illness.^**17**^ In addition, Ian and colleagues, who investigated the association of inflammatory biomarkers with the COVID-19 outcome, have found that the elevated PCT was associated with increased severity and mortality of the disease but not with an increased need for ICU admission.^**18**^

In order to monitor the evolution of the PCT level and measure its relative changes to the initial value, we calculated the PCT-c. We observed that its low levels were significantly associated with COVID-19 in-hospital mortality (p < 0.01). Whereas during follow-up, it was observed to have progressively decreased in non-survivors and increased in survivors. Confusingly, PCT-c D3 at the cutoff of -11.1% showed the lowest AUC (0.74), specificity (87.9%), PPV (71.4%) and accuracy (83.0%). This could be explained by the small drop in some PCTD3 values in non-survivors, with a small rise in some survivors. While it is difficult to assess the impact of not specifically studied variables, we attributed this to the varying timing of therapy initiation.

Ruiz-Rodríguez et al. and Suberviola et al. have introduced PCT-c to assess the utility of serial PCT and its relationship with mortality.^**7**,**19**^ Ruiz-Rodriguez and colleagues determined the clearance of PCT after 24, 48, and 72 hours of treatment of 27 patients with septic shock and found a significant increase in PCT-c in survivors compared to non-survivors.^**7**^ Meanwhile, Suberviola and co-workers studied 88 patients with septic shock admitted to the ICU and found that the mortality in patients with increased PCT-c in the first 72 hours of treatment was significantly lower than in patients with reduced clearance in the same period (15.4% versus 58.8%, p <0.01).^**19**^

Consistent with the fact that cytokines released in COVID-19 infection, particularly (INF)-γ, have a negative effect on PCT levels,^**20**^ Schuetz has recently suggested PCT as a valuable tool in identifying COVID-19 patients at high risk for clinical deterioration. Furthermore, he stated that at the time of hospital admission, most COVID-19 patients showed very low PCT levels (< 0.25 or even < 0.1ng/ml). However, in clinical deterioration of mild cases, which were expected to have low PCT levels, significant progressive elevation in their level occurred, which added to the strength of the PCT prognostic ability.^**21**^

Several studies also reported that levels of PCT in COVID-19 patients upon hospital admission were typically normal, as in other viral infections, and increased afterward in patients admitted to the ICU.^**2**,**15**, **22**^ PCT elevation in these cases was attributed to bacterial co-infection (lung damage by the virus gave access to normal bacterial flora which became invasive and developed secondary bacterial pneumonia), or, on the other hand, patient deterioration with the advancement of the hyperinflammatory syndrome and cytokine storm (increased synthesis of PCT by cytokines associated with immune dysregulation could develop severe inflammatory pneumonitis and endothelial dysfunction).^**21**,**23**^

In concordance with these data, our results showed that serial PCT and PCT-c had an overall better predictive performance than initial PCT for in-hospital mortality of COVID-19 patients. Where, at optimum cut-off values, the AUC followed the order of PCTD5 > PCT-c D5 > PCTD3 > PCTD1 > PCT-c D3 being 0.96, 0.89, 0.82, 0.81 and 0.74, respectively. In addition, the combined prediction value was better than the single prediction by the initial PCT. At optimum cut-off values, the AUC increased when PCTD1 was added to PCTD3, PCTD5, PCT-cD3, or PCT-cD5 to be 0.93, 0.98, 0.96, and 1.00, respectively.

One disadvantage of serial PCT measurement could be the cost. However, the overall cost of aggressive unnecessary therapeutic interventions and the resulting patients’ complications could exceed the cost of the repetition of the test. To our knowledge, our study is the first to explore the role of PCT-c as an indicator for dynamic changes in PCT to predict the outcome of hospitalized COVID-19 patients, but it has several limitations. The first issue is the small sample size and the disproportionate number of cases in the study groups. Second, we did not collect enough information about the antibiotic treatment of patients or the results of their microbiological cultures. Third, we studied changes in PCT alone and did not account for the possibility that the relationship of PCT to other inflammatory markers could be more informative. Thus, further studies on larger samples and more clinical considerations and correlation and comparison with other inflammatory markers are required.

## Conclusions

Persistently high PCT concentrations and reduced PCT-c were associated with significantly higher COVID-19 in-hospital mortality. This suggests that the elevated PCT, with its progressive kinetics, may help predict the outcomes of COVID-19 patients. Moreover, PCT-c can be used to assess PCT kinetic changes during the disease course and evaluate the potential value of PCT as a prognostic marker.

## Data Availability

All data are presented in the main manuscript

## List of abbreviations

COVID-19: Coronavirus disease 2019
INF-γ: Interferon-gamma
PCT: Procalcitonin
PCT-c: Procalcitonin clearance
RT-PCR: Reverse transcription-polymerase chain reaction
SARS-CoV-2: Severe acute respiratory syndrome corona virus 2
TNF-α: Tumor necrosis factor-alpha

## Acknowledgement

None.

## Authors’ contribution

All authors contributed significantly to this work, whether it was in the conception, study design, data acquisition, analysis, and interpretation, or in the drafting, revising, or reviewing of the article, and they all provided final approval of the version to be published.

## Data availability statement

On reasonable request, the corresponding author will provide the datasets used and/or analyzed during the current work.

## Ethics

Ethical approval for the current study protocol was obtained from Ain Shams University Faculty of Medicine Research Ethics Committee (REC) FWA 00017585.

## Statement of Informed Consent

All procedures were explained to all participants or their first-degree relatives, with informed consents obtained from them.

## Consent for publication

Not applicable.

## References

1. Xu J, Xu C, Zhang R, et al (2020) Associations of procalcitonin, C-reaction protein and neutrophil-to-lymphocyte ratio with mortality in hospitalized COVID-19 patients in China. Scientific Reports, 10(1):15058.

2. Chen N, Zhou M, Dong X, et al (2020) Epidemiological and clinical characteristics of 99 cases of 2019 novel coronavirus pneumonia in Wuhan, China: a descriptive study. The Lancet, 395(10223):507–513.

3. Armstrong RA, Kane AD, Cook TM (2020) Outcomes from intensive care in patients with COVID-19: a systematic review and meta-analysis of observational studies. Anaesthesia 2020; 75(10):1340–1349.

4. Ahmed S, Siddiqui I, Jafri L, et al (2018) Prospective evaluation of serum procalcitonin in critically ill patients with suspected sepsis-experience from a tertiary care hospital in Pakistan. Annals of Medicine and Surgery, 35:180–184.

5. Azevedo JR, Torres OJ, Czeczko NG, et al (2012). Procalcitonina como biomarcador de prognóstico da sepse grave e choque séptico. Revista do Colégio Brasileiro de Cirurgiões, 39(6):456–461.

6. Badawy Abdelfattah E, Ahmed El-Zahapy H, El Said A, et al (2020) Hospital response to COVID-19 a consensus: Report on Ain Shams University Hospital Strategy. ScienceOpen [published online ahead of print].

7. Ruiz-Rodríguez JC, Caballero J, Ruiz-Sanmartin A, et al (2012) Usefulness of procalcitonin clearance as a prognostic biomarker in septic shock. A prospective pilot study. Medicina Intensiva (English Edition), 36(7):475–480.

8. Assicot M, Gendrel D, Carsin H, et al (1993) High serum procalcitonin concentrations in patients with sepsis and infection. The Lancet, 341(8844):515–518.

9. Müller B, Becker KL, Schächinger H, et al (2000) Calcitonin precursors are reliable markers of sepsis in a medical intensive care unit. Critical Care Medicine, 28(4):977– 983.

10. Schuetz P, Wirz Y, Sager R, et al (2017). Procalcitonin to initiate or discontinue antibiotics in acute respiratory tract infections. Cochrane Database of Systematic Reviews. 10(10):CD007498.

11. Bhandari S, Bhargava A, Sharma S, et al. (2020) Clinical Profile of Covid-19 Infected Patients Admitted in a Tertiary Care Hospital in North India. The Journal of the Association of Physicians of India. 68(5):13–17.

12. Cao H, Ruan L, Liu J, et al (2020) The clinical characteristic of eight patients of COVID-19 with positive RT-PCR test after discharge. Journal of Medical Virology. 92(10):2159–2164.

13. Sun D, Li H, Lu XX, et al. (2020) Clinical features of severe pediatric patients with coronavirus disease 2019 in Wuhan: a single center’s observational study. World Journal of Pediatrics. 16(3):251–259.

14. Zhang JJ, Dong X, Cao YY, et al (2020) Clinical characteristics of 140 patients infected with SARS-CoV-2 in Wuhan, China. Allergy. 75(7):1730–1741

15. Huang C, Wang Y, Li X, et al (2020) Clinical features of patients infected with 2019 novel coronavirus in Wuhan, China. Lancet. 395(10223):497–506.

16. Shi S, Qin M, Shen B, et al (2020) Association of cardiac injury with mortality in hospitalized patients with COVID-19 in Wuhan, China. JAMA Cardiology. 5(7):802–810.

17. Lippi G, Plebani M, Henry BM (2020) Thrombocytopenia is associated with severe coronavirus disease 2019 (COVID-19) infections: A meta-analysis. Clinica Chimica Acta. 506:145–148.

18. Huang I, Pranata R, Lim MA, et al (2020) C-reactive protein, procalcitonin, D-dimer, and ferritin in severe coronavirus disease-2019: a meta-analysis. Therapeutic Advances in Respiratory Disease. 14:1753466620937175.

19. Suberviola B, Castellanos-Ortega A, González-Castro A, et al (2012) Prognostic value of procalcitonin, C-reactive protein and leukocytes in septic shock. Medicina Intensiva. 36:177–184.

20. Cleland DA and Eranki AP (2021) Procalcitonin. [online] PubMed. Available at: https://www.ncbi.nlm.nih.gov/books/NBK539794/. [Accessed 10 Jul. 2021].

21. Schuetz P (2020) The role of procalcitonin for risk assessment and treatment of COVID-19 patients. [online] HealthManagement. Available at: https://healthmanagement.org/c/healthmanagement/issuearticle/the-role-of-procalcitonin-for-risk-assessment-and-treatment-of-covid-19-patients [Accessed 10 Jul. 2021].

22. Wang D, Hu B, Hu C, et al (2020) Clinical characteristics of 138 hospitalized patients with 2019 novel coronavirus–infected pneumonia in Wuhan, China. JAMA. 323(11):1061–1069

23. Shah V (2021) Meaning of elevated procalcitonin unclear in COVID-19. [online] Massachusetts General Hospital: Advances in Motion. Available at: https://advances.massgeneral.org/researchand-innovation/article.aspx?id=1174 [accessed 23 May 2021].

